# Defining reference values for body composition indexes by magnetic resonance imaging in UK Biobank

**DOI:** 10.1101/2022.06.22.22276739

**Authors:** Liang Dai, Xiao-yan Huang, Yue-qi Lu, Yu-yang Liu, Cong-ying Song, Jing-wen Zhang, Jing Li, Yue Zhang, Ying Shan, Yu Shi

## Abstract

**Background:** Magnetic resonance imaging (MRI) is the gold standard for body composition measurements, while such reference range data have not been reported yet.

**Methods:** We used MRI-measured body composition data from the UK Biobank cohort, including three lean tissue parameters and seven adipose tissue parameters. Participants aged between 45 to 74 years from both genders with at least one parameter data were screened. Age- and sex-specific percentile curves were generated by the lambda–mu–sigma method. Three levels of reference ranges were provided, which were equivalent to mean ± 1 standard deviation (SD), 2 SDs, and 2.5 SDs, respectively.

**Results:** The final analysis set for each parameter ranged from 4 842 to 14 148 participants (53.4-56.6 % women) with median ages around 61 years after eligibility confirmation. For lean tissue parameters, from age 45 to age 70, median percentiles of total lean tissue volume and total thigh fat-free muscle volume reduced by 2.83 L and 1.73 L in men, and by 3.02 L and 1.51 L in women, respectively. Within the age range, median percentiles of weight-to-muscle ratio elevated by 0.51 kg/L in men and 0.83 kg/L in women, respectively. Adipose tissue parameters presented inconsistent trajectories. For men, median percentiles of muscle fat infiltration, visceral adipose tissue (VAT) volume, total abdominal adipose tissue index, and abdominal fat ratio elevated by 1.48 %, 0.32 L, 0.08 L/m^2^ and 0.04, while abdominal subcutaneous adipose tissue (ASAT) volume and total adipose tissue volume reduced by 0.47 L and 0.41 L from age 45 to age 70, respectively. And total trunk fat volume remained stable with aging (approximate 9.52 L). For women, muscle fat infiltration and VAT volume showed similar trends with men, increased by 1.68 % and 0.76 L from age 45 to age 70. Other five parameters firstly increased from age 45 to age 60 (ASAT volume: 0.35 L, total adipose tissue volume: 0.78 L, total trunk fat volume: 1.12 L, total abdominal adipose tissue index: 0.49 L/m^2^, and abdominal fat ratio: 0.06), then kept stable (total abdominal adipose tissue index and abdominal fat ratio) or presented a slight reduction afterwards (ASAT volume: 0.33 L, total adipose tissue volume: 0.14 L, and total trunk fat volume: 0.20 L).

**Conclusions:** We have established reference ranges for MRI-measured body composition parameters based on large community-dwelling population. The data would benefit more accurate assessment of abnormal adipose and muscle conditions.

## Introduction

Both lean mass and fat mass of the human body are critical for various health-related outcomes. It has been illustrated that a U-shaped relation exists between lean body mass and all-cause mortality [1]. For another, the intense relationship has also been repeatedly proven between adipose tissue and cardio-metabolic health [2, 3]. Hence, body composition evaluations benefit both clinicians and patients in understanding potential risks of negative health events. Body mass index (BMI), the classical body measurement index, has been criticized for a long time due to its poor accuracy for specific body composition [4, 5]. Waist circumstance (WC) and waist-hip ratio (WHR), served as more sensitive indicators for adipose tissue accumulation, cannot comprehensively interpret potential disease risks [6, 7]. Therefore, more comprehensive body composition measurements are encouraged in clinical practice.

Bioelectrical impedance analysis (BIA) and dual-energy X-ray absorptiometry (DXA) are two commonly-used instruments for body composition analysis. BIA can estimate the adipose and muscle mass on the foundation of whole-body electrical conductivity [8]. Due to its convenience and low-cost, BIA is frequently used for body composition screening. DXA, as the name suggests, calculates body tissue mass based on absorption and depletion differences of X-rays with two energies [9]. As a snapshot technique, DXA can provide composition indexes in specific limbs. However, both instruments have their own limitations, such as inconsistent results from different brands, interference from body hydration status, and incapability to assess intra-muscular fat, which would lead to obesity paradox or muscle quality overestimation [10-13, S1-2]. It is a certain trend that more accurate instrument is needed for body composition evaluation.

Magnetic resonance imaging (MRI) technique utilizes the interaction of hydrogen nuclei with magnetic field of the instrument to distinguish different body components [14], and serves as the gold standard for body composition measurements [15]. Owing to technology update, difficulties such as anatomical level selection and imaging analysis of MRI instrument have been much overcome [16]. In addition, no radiation exposure and reliable reproducibility further prompt MRI to be a promising technique in body composition assessment [17]. Currently no reference range data of body composition parameters assessed by MRI technique are reported, while interestingly, such studies have been published regarding BIA and DXA [18-19, S3]. Hence, we conducted this study to fill this gap. By using data from the UK Biobank, we developed sex-specific reference ranges for body composition indexes in a large community-dwelling population of middle-aged adults (aged 45-59 years) and young elders (aged 60-74 years), hoping to supplement MRI-based body composition standard for future studies.

## Methods

### Participants

The UK Biobank, initiated in 2006, has recruited more than half a million volunteers aged 40 to 69 years old via the United Kingdom National Health Service registers between April 2007 and December 2010. After baseline assessment, an imaging visit was arranged for approximate ten thousand participants since 2014, covering brain, heart, and body composition MRI measurements [20]. The UK Biobank got ethical approval from the North West Multi-Center Research Ethics Committee (REC reference: 11/NW/03820) and all participants provided written informed consent. The corresponding research plan for this study had been submitted to the UK Biobank (application ID 65814).

Our study screened subjects with MRI-measured body composition data from the UK Biobank cohort. Ten parameters were analyzed, including three lean tissue parameters (total lean tissue volume, total thigh fat-free muscle volume (FFMV), and weight-to-muscle ratio), and seven adipose tissue parameters (muscle fat infiltration, abdominal subcutaneous adipose tissue (ASAT) volume, visceral adipose tissue (VAT) volume, total trunk fat volume, total adipose tissue volume, total abdominal adipose tissue index, and abdominal fat ratio). Participants aged between 45 to 74 from both genders and possessed with data for at least one parameter mentioned above were included. Ethnic minority, defined as sample size cannot support statistical estimation [21], and objective measurement mistakes based on documented error indicators were excluded from the analysis sets. We also excluded diseases, combined medications, surgeries and other physiological or pathological conditions that would influence the distribution of lean and fat mass, including endocrine, nutritional and metabolic diseases (ICD-10: E00-07, E10-14, E15-16, E20-E32, E34-35, E40-46, E50-56, E58-61, E63, E64, E70-80, E83-90), neoplasms (ICD-10: C00-97, D00-09, D37-38), human immunodeficiency virus infection (ICD-10: B20-24), administration of anti-obesity preparations, systemic hormonal preparations and anti-diabetes products, pregnancy, and certain abdominal wall, subcutaneous tissue and miscellaneous surgeries (OPCS3: 400, 878-879, 928; OPCS4: S50-52, S62.2-62.9, S63.8-63.9, T32, T76-77, X02.1, X02.8-X02.9, X05, X09.1, X09.3, X09.8-09.9, X15, X22). The detailed participants’ flowchart for each parameter is shown in File S1.

### MRI protocol and body composition analysis

MRI procedure was conducted using a Siemens MAGNETOM Aera 1.5 T scanner (Siemens Healthineers, Erlangen, Germany) with the dual-echo Dixon Vibe protocol from neck to knees. The imaging provided water and fat separated volumetric data set for body composition analysis. Detailed instrument process and analysis methods have been published elsewhere [22-24, S4]. In brief, the total 1.1m scanning was divided into six overlapping slabs, with 64 slices in slabs 1 and 6, 44 slices in slabs 2 to 4, and 72 slices in slab 5. The obtained images went through calibration, stacks fusion, and segmentation. After professional inspection by analysis engineer, lean and adipose volumes were finally determined [25]. The body composition analyses were completed by AMRA Profiler™ (AMRA AB, Linköping, Sweden).

### Statistical analysis

Enrolled participants’ basic characteristics and body composition parameters are presented as mean (standard deviation), or median (interquartile range) based on data features. Considering that body composition varies between two genders, we provided the sex-specific reference ranges.

We generated sex-specific percentile curves and age-and-sex-specific reference ranges using the lambda–mu–sigma (LMS) method [26]. The LMS method assumes underlying frequency distributions of the data, and estimates each of the distribution moments in the form of smooth curves plotted against age. Box-Cox Cole and Green (BCCG), Box-Cox power exponential (BCPE) and Box-Cox t (BCT) are LMS class of distributions. BCCG model estimates the first three moments of the measurement distribution as the age-varying median (μ), the coefficient of variation (σ), and skewness in the form of a Box-Cox transformation (λ). BCPE and BCT models extend BCCG model by adjusting for kurtosis (τ), based respectively on the power exponential distribution and the t distribution. In this study, for each body composition parameter, we fitted the three models and selected the optimal one based on local maximum likelihood. We generated reference curves and provided reference ranges for both genders at specific ages according to the estimated coefficients in the optimal model. The R package “gamlss” was used for the above analysis.

In addition, according to the age segmentation criteria [27], we divided the whole population into two sub-categories, namely middle-aged adults (aged 45-59 years) and young elders (aged 60-74 years). Then, we conducted a consistency test (via Student’s t test) to detect if the measurement values in the sub-categories had significant difference, thus, to determine whether the reference ranges should be presented separately or jointly. The reference ranges were presented as mean ± 1 standard deviation (SD), mean ± 2SD, and mean ± 2.5SD.

## Results

### Participants’ characteristics

The characteristics of included participants are shown in *Table* 1. The number of participants varied for ten parameters; hence the data analysis sets of the body composition were also different accordingly. The participant flowchart can be found in *Figure* S1. Because of low statistical power of ethnic minority, only Caucasians were included in further analyses. After confirmation of prescribed eligible criteria, the final data sets for ten parameters ranged from 4 842 to 14 148.

**Table 1.** Participants’ characteristics

| Parameter | Gender | Median (IQR) | Age (mean±SD,<br>years) | Height (mean±SD,<br>m) | Weight (median (IQR),<br>kg) | BMI (median (IQR),<br>kg/m<sup>2</sup>) |
| --- | --- | --- | --- | --- | --- | --- |
| Total lean tissue volume (L) | Male (n=2 122) | 28.37 (26.28, 30.67) | 60.63 ± 7.21 | 1.76 ± 0.06 | 81.20 (73.50, 89.00) | 26.13 (24.03, 28.45) |
|  | Female (n=2 720) | 20.51 (18.94, 22.26) | 60.23 ± 7.10 | 1.63 ± 0.06 | 66.50 (59.90, 75.00) | 25.00 (22.61, 28.08) |
| Total thigh FFMV (L) | Male (n=5 697) | 12.41 (11.33, 13.55) | 61.28 ± 7.06 | 1.76 ± 0.06 | 80.80 (73.50, 89.20) | 26.04 (23.92, 28.51) |
|  | Female (n=7 416) | 8.19 (7.45, 8.96) | 60.92 ± 6.86 | 1.63 ± 0.06 | 66.50 (59.60, 74.90) | 24.95 (22.55, 27.99) |
| Weight-to-muscle ratio (kg/L) | Male (n=5 697) | 6.49 (6.05, 7.03) | 61.28 ± 7.06 | 1.76 ± 0.06 | 80.80 (73.50, 89.20) | 26.04 (23.92, 28.51) |
|  | Female (n=7 415) | 8.13 (7.44, 8.93) | 60.93 ± 6.86 | 1.63 ± 0.06 | 66.50 (59.60, 74.90) | 24.95 (22.55, 28.00) |
| Muscle fat infiltration (%) | Male (n=5 674) | 6.15 (5.33, 7.20) | 61.28 ± 7.05 | 1.76 ± 0.06 | 80.80 (73.50, 89.20) | 26.05 (23.92, 28.52) |
|  | Female (n=7 388) | 7.30 (6.37, 8.44) | 60.91 ± 6.86 | 1.63 ± 0.06 | 66.50 (59.60, 74.90) | 24.95 (22.54, 27.98) |
| ASAT volume (L) | Male (n=6 544) | 5.23 (4.05, 6.74) | 61.12 ± 7.06 | 1.77 ± 0.07 | 81.80 (74.40, 90.60) | 26.20 (24.05, 28.73) |
|  | Female (n=7 493) | 7.22 (5.37, 9.50) | 60.94 ± 6.86 | 1.63 ± 0.06 | 66.50 (59.60, 74.80) | 24.94 (22.54, 27.96) |
| VAT volume(L) | Male (n=6 574) | 4.39 (2.92, 6.00) | 61.12 ± 7.06 | 1.77 ± 0.07 | 81.90 (74.40, 90.80) | 26.22 (24.07, 28.76) |
|  | Female (n=7 574) | 2.20 (1.39, 3.25) | 60.93 ± 6.87 | 1.63 ± 0.06 | 66.60 (59.70, 75.05) | 24.97 (22.58, 28.09) |
| Total abdominal adipose tissue index (L/m<sup>2</sup>) | Male (n=6 539) | 3.13 (2.31, 4.03) | 61.11 ± 7.06 | 1.77 ± 0.07 | 81.80 (74.40, 90.60) | 26.20 (24.05, 28.73) |
|  | Female (n=7 487) | 3.59 (2.59, 4.78) | 60.94 ± 6.87 | 1.63 ± 0.06 | 66.50 (59.60, 74.80) | 24.94 (22.54, 27.96) |
| Total adipose tissue volume (L) | Male (n=2 122) | 18.10 (14.48, 22.25) | 60.63 ± 7.21 | 1.76 ± 0.06 | 81.20 (73.50, 89.00) | 26.13 (24.03, 28.45) |
|  | Female (n=2 720) | 20.30 (16.18, 25.37) | 60.23 ± 7.10 | 1.63 ± 0.06 | 66.50 (59.90, 75.00) | 25.00 (22.61, 28.08) |
| Total trunk fat volume (L) | Male (n=5 668) | 9.58 (7.09, 12.31) | 61.28 ± 7.05 | 1.76 ± 0.06 | 80.80 (73.50, 89.10) | 26.03 (23.92, 28.50) |
|  | Female (n=7 329) | 9.53 (6.90, 12.62) | 60.92 ± 6.86 | 1.63 ± 0.06 | 66.40 (59.60, 74.60) | 24.92 (22.52, 27.89) |
| Abdominal fat ratio | Male (n=5 668) | 0.44 (0.37, 0.50) | 61.28 ± 7.05 | 1.76 ± 0.06 | 80.80 (73.50, 89.10) | 26.03 (23.92, 28.50) |
|  | Female (n=7 329) | 0.54 (0.46, 0.61) | 60.92 ± 6.86 | 1.63 ± 0.06 | 66.40 (59.60, 74.60) | 24.92 (22.52, 27.89) |

ASAT, abdominal subcutaneous adipose tissue; FFMV, fat-free muscle volume; IQR, interquartile range; SD, standard deviation; VAT, visceral adipose tissue.

For the three lean volume parameters, medians of total lean tissue volume and total thigh FFMV were higher in men than in women, while median weight-to-muscle ratio was lower. For the seven fat volume parameters, median muscle fat infiltration, ASAT volume, total adipose tissue volume, total abdominal adipose tissue index, and abdominal fat ratio were higher in women than in men, but median VAT volume was higher in men compared with women. The remaining total trunk fat volume was comparable between women and men.

### Lean volume parameters

Reference percentiles of lean tissue parameters are shown in *Table* 2. Both total lean tissue volume and total thigh FFMV showed a steady downward trend in either gender (*Figure* 1). For men, median total lean tissue volume and total thigh FFMV decreased by 2.83 L and 1.73 L from age 45 to age 70, respectively. Similarly, the reductions of these two parameters were 3.02 L and 1.51 L in women from age 45 to age 70, respectively. Weight-to-muscle ratio presented a gradual rising trend in both genders (*Figure* 1). Compared with age 45, median percentiles of weight-to-muscle ratio increased by 0.51 kg/L in men and 0.83 kg/L in women at age 70, respectively.

**Table 2.** Reference percentiles of lean tissue parameters for both genders at specific ages

|  | Age<br>(years) | Total lean tissue volume<br>(L) | Total thigh FFMV<br>(L) | Weight-to-muscle ratio<br>(kg/L) |
| --- | --- | --- | --- | --- |
| Male | 45 | 30.14 (24.52, 38.19) | 13.53 (10.34, 17.27) | 6.19 (5.04, 7.84) |
|  | 50 | 29.62 (23.78, 36.75) | 13.23 (10.14, 16.80) | 6.28 (5.14, 7.91) |
|  | 55 | 29.07 (23.09, 35.60) | 12.91 (9.93, 16.31) | 6.37 (5.23, 8.01) |
|  | 60 | 28.50 (22.62, 34.73) | 12.57 (9.69, 15.79) | 6.47 (5.33, 8.12) |
|  | 65 | 27.91 (22.22, 33.94) | 12.19 (9.42, 15.24) | 6.58 (5.43, 8.27) |
|  | 70 | 27.31 (21.69, 33.01) | 11.80 (9.14, 14.69) | 6.70 (5.53, 8.44) |
| Female | 45 | 22.75 (18.26, 28.20) | 9.32 (7.13, 12.22) | 7.47 (5.79, 9.85) |
|  | 50 | 21.78 (17.19, 27.43) | 8.83 (6.80, 11.43) | 7.77 (6.05, 10.27) |
|  | 55 | 20.97 (16.57, 26.41) | 8.44 (6.53, 10.81) | 8.04 (6.26, 10.66) |
|  | 60 | 20.45 (16.43, 25.33) | 8.19 (6.35, 10.39) | 8.18 (6.39, 10.87) |
|  | 65 | 20.06 (16.27, 24.64) | 8.00 (6.21, 10.10) | 8.23 (6.48, 10.88) |
|  | 70 | 19.73 (15.96, 24.29) | 7.81 (6.05, 9.84) | 8.30 (6.58, 10.91) |

FFMV, fat-free muscle volume.
Reference percentile was defined as median (3rd, 97th).

**Figure 1.**
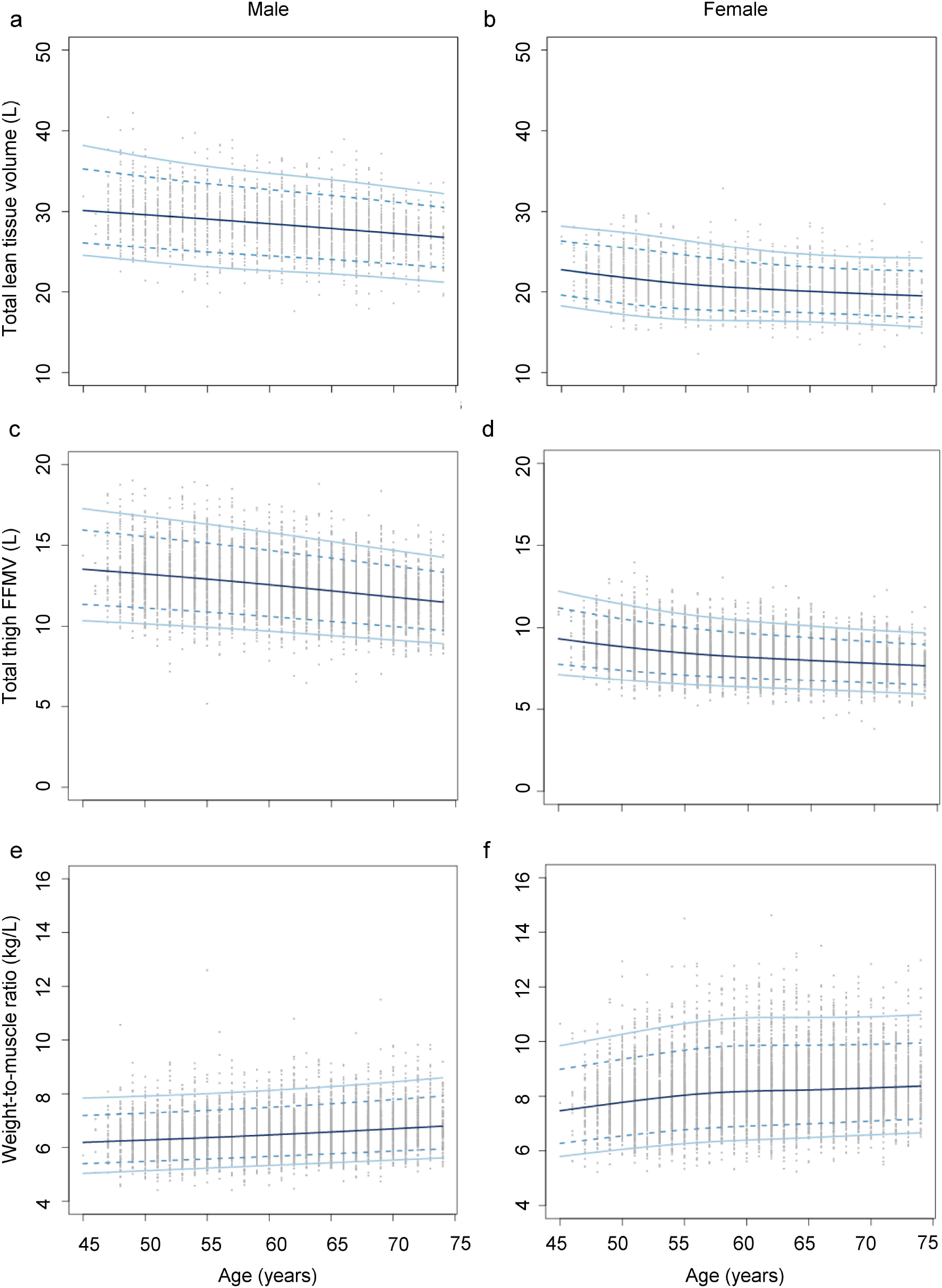
Age-specific and sex-specific percentile curves for (*a*) total lean tissue volume in male, (*b*) total lean tissue volume in female, (*c*) total thigh fat-free muscle volume (FFMV) in male, (*d*) total thigh FFMV in female, (*e*) weight-to-muscle ratio in male, and (*f*) weight-to-muscle ratio in female.

Reference data of lean volume parameters for middle-aged adults and young elders are presented in *Table* 3. Three levels of ranges were given, which were equivalent to mean ± 1 standard deviation (SD), mean ± 2 SDs, and mean ± 2.5 SDs. The consistency tests between two populations were conducted in both genders and significant difference was found in each parameter (*Table* S1), which indicated that the reference ranges should be considered independently for middle-aged adults and young elders.

**Table 3.** Reference data of middle-aged adults and young elders for lean tissue parameters

|  |  | Male |  |  | Female |  |  |
| --- | --- | --- | --- | --- | --- | --- | --- |
|  | Parameters | Mean±SD | Mean±2SD | Mean±2.5SD | Mean±SD | Mean±2SD | Mean±2.5SD |
| Middle-aged adults (45-59 years) | Total lean tissue volume (L) | [25.98, 32.83] | [22.55, 36.26] | [20.83, 37.98] | [18.65, 24.09] | [15.92, 26.82] | [14.56, 28.18] |
|  | Total thigh FFMV (L) | [11.33, 14.76] | [9.61, 16.48] | [8.75, 17.34] | [7.40, 9.78] | [6.21, 10.97] | [5.62, 11.57] |
|  | Weight-to-muscle ratio (kg/L) | [5.67, 7.17] | [4.92, 7.92] | [4.54, 8.29] | [6.91, 9.27] | [5.73, 10.45] | [5.14, 11.04] |
| Young elders (60-74 years) | Total lean tissue volume (L) | [24.71, 30.93] | [21.60, 34.04] | [20.04, 35.60] | [17.90, 22.40] | [15.65, 24.65] | [14.53, 25.78] |
|  | Total thigh FFMV (L) | [10.57, 13.68] | [9.01, 15.24] | [8.23, 16.02] | [6.97, 9.06] | [5.93, 10.10] | [5.40, 10.63] |
|  | Weight-to-muscle ratio (kg/L) | [5.92, 7.46] | [5.15, 8.23] | [4.77, 8.61] | [7.17, 9.58] | [5.97, 10.78] | [5.37, 11.38] |

FFMV, fat-free muscle volume; SD, standard deviation.

Reference values used in LMS method for lean volume parameters are available in Data S1.

### Fat volume parameters

Reference percentiles of adipose tissue parameters are shown in *Table* 4. For men, four parameters (muscle fat infiltration, VAT volume, total abdominal adipose tissue index, and abdominal fat ratio) presented slight increasing trend with aging, two parameters (ASAT volume and total adipose tissue volume) presented gradual decreasing trend, and one parameter (total trunk fat volume) remained stable (*Figures* 2 and S1). Compared with age 45, median percentiles of muscle fat infiltration, VAT volume, total abdominal adipose tissue index, and abdominal fat ratio elevated by 1.48 %, 0.32 L, 0.08 L/m^2^, and 0.04 at age 70, respectively. Median percentiles of ASAT volume and total adipose tissue volume reduced by 0.47 L and 0.41 L from age 45 to age 70, respectively. For women, only muscle fat infiltration and VAT volume showed similar trends with men, increasing from 6.25 % and 1.61 L at age 45 to 7.93 % and 2.37 L at age 70, respectively (*Figure* 3). Total abdominal adipose tissue index and abdominal fat ratio firstly increased from age 45 to age 60 (3.16 L/m^2^ to 3.65 L/m^2^, and 0.49 to 0.55, respectively), then kept stable afterwards (*Figures* 3 and S1). The remaining three parameters, ASAT volume, total adipose tissue volume, and total trunk fat volume, presented rising trend from age 45 to age 60 at first, then a slight reduction after age 60. To be specific, compared with age 45, ASAT volume, total adipose tissue volume, and total trunk fat volume increased by 0.35 L, 0.78 L, and 1.12 L, respectively. Then, they decreased by 0.33 L, 0.14 L, and 0.20 L from age 60 to age 70, respectively.

**Table 4.** Reference percentiles of fat tissue parameters for both genders at specific ages

|  | Age | Muscle fat |  |  | Total abdominal |  |  |  |
| --- | --- | --- | --- | --- | --- | --- | --- | --- |
|  | (years) | infiltration (%) | ASAT volume (L) | VAT volume (L) | adipose tissue index<br>(L/m<sup>2</sup>) | Total adipose tissue<br>volume (L) | Total trunk fat<br>volume (L) | Abdominal fat<br>ratio |
| Male | 45 | 5.25 (3.74, 8.09) | 5.55 (2.14, 12.52) | 4.09 (0.96, 9.25) | 3.06 (0.98, 6.38) | 18.41 (7.20, 33.07) | 9.52 (3.27, 19.59)) | 0.41 (0.19, 0.58) |
|  | 50 | 5.49 (3.91, 8.49) | 5.47 (2.17, 12.02) | 4.17 (1.04, 9.25) | 3.08 (1.05, 6.30) | 18.31 (7.83, 32.61) | 9.52 (3.35, 19.30) | 0.42 (0.20, 0.59) |
|  | 55 | 5.76 (4.09, 8.93) | 5.37 (2.21, 11.54) | 4.24 (1.12, 9.25) | 3.10 (1.11, 6.22) | 18.22 (8.34, 32.20) | 9.53 (3.43, 19.02) | 0.43 (0.21, 0.59) |
|  | 60 | 6.05 (4.29, 9.42) | 5.28 (2.23, 11.09) | 4.30 (1.19, 9.26) | 3.11 (1.15, 6.15) | 18.14 (8.76, 31.82) | 9.53 (3.51, 18.74) | 0.43 (0.22, 0.59) |
|  | 65 | 6.38 (4.51, 9.96) | 5.18 (2.24, 10.66) | 4.36 (1.25, 9.28) | 3.12 (1.19, 6.06) | 18.06 (9.11, 31.47) | 9.53 (3.60, 18.45) | 0.44 (0.23, 0.60) |
|  | 70 | 6.73 (4.75, 10.56) | 5.08 (2.25, 10.26) | 4.41 (1.30, 9.31) | 3.14 (1.20, 5.96) | 18.00 (9.41, 31.15) | 9.53 (3.69, 18.17) | 0.45 (0.24, 0.60) |
| Female | 45 | 6.25 (4.08, 10.09) | 7.05 (2.55, 15.33) | 1.61 (0.43, 5.06) | 3.16 (1.10, 7.19) | 19.70 (9.43, 38.92) | 8.60 (2.93, 20.28) | 0.49 (0.25, 0.66) |
|  | 50 | 6.57 (4.58, 9.86) | 7.19 (2.64, 15.24) | 1.85 (0.49, 5.35) | 3.33 (1.18, 7.32) | 20.09 (9.81, 38.53) | 9.02 (3.14, 20.14) | 0.51 (0.27, 0.67) |
|  | 55 | 6.90 (4.88, 10.25) | 7.35 (2.75, 15.24) | 2.08 (0.55, 5.63) | 3.53 (1.26, 7.52) | 20.39 (10.14, 38.13) | 9.47 (3.36, 20.22) | 0.53 (0.29, 0.69) |
|  | 60 | 7.24 (5.03, 11.10) | 7.40 (2.81, 15.03) | 2.24 (0.60, 5.73) | 3.65 (1.32, 7.58) | 20.48 (10.35, 37.47) | 9.72 (3.52, 19.99) | 0.55 (0.30, 0.70) |
|  | 65 | 7.58 (5.38, 11.40) | 7.25 (2.81, 14.41) | 2.32 (0.62, 5.67) | 3.66 (1.34, 7.39) | 20.41 (10.47, 36.61) | 9.65 (3.56, 19.21) | 0.55 (0.31, 0.69) |
|  | 70 | 7.93 (5.61, 12.10) | 7.07 (2.80, 13.73) | 2.37 (0.63, 5.57) | 3.64 (1.36, 7.16) | 20.34 (10.58, 35.85) | 9.52 (3.58, 18.42) | 0.55 (0.33, 0.69) |

ASAT, abdominal subcutaneous adipose tissue; VAT, visceral adipose tissue.
Reference percentile was defined as median (3rd, 97th).

**Table 5.**
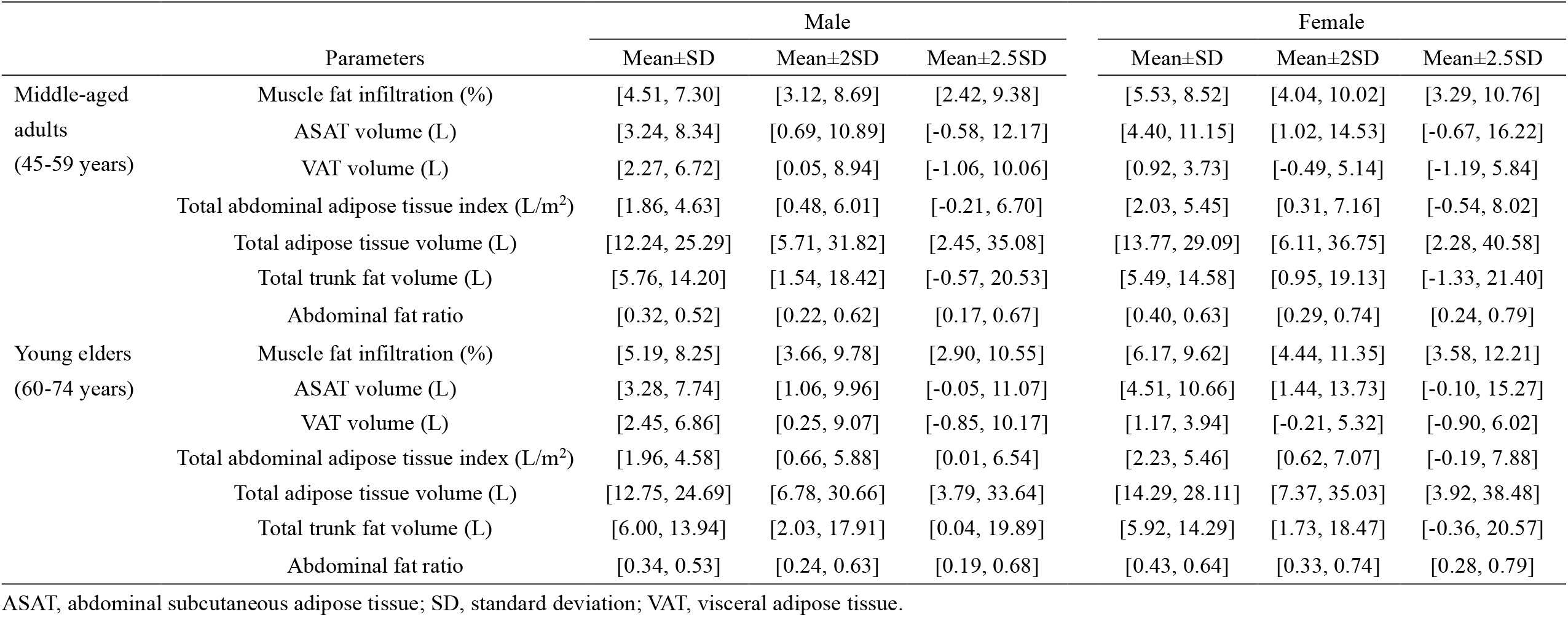
Reference data of middle-aged adults and young elders for fat tissue parameters

**Figure 2.**
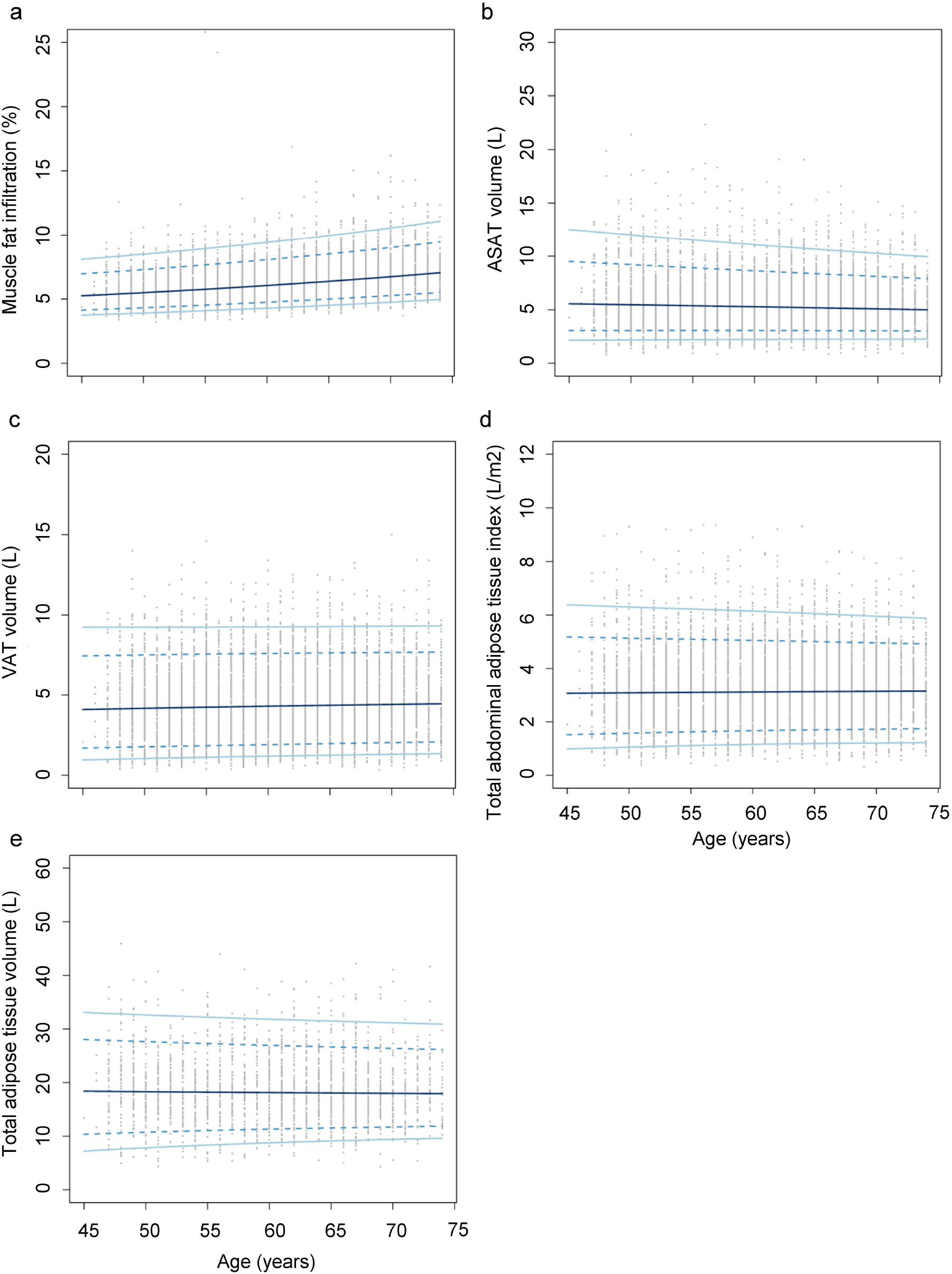
Age-specific and sex-specific percentile curves for (*a*) muscle fat infiltration, (*b*) abdominal subcutaneous adipose tissue (ASAT) volume, (*c*) visceral adipose tissue (VAT) volume, (*d*) total abdominal adipose tissue index, and (*e*) total adipose tissue volume in male.

**Figure 3.**
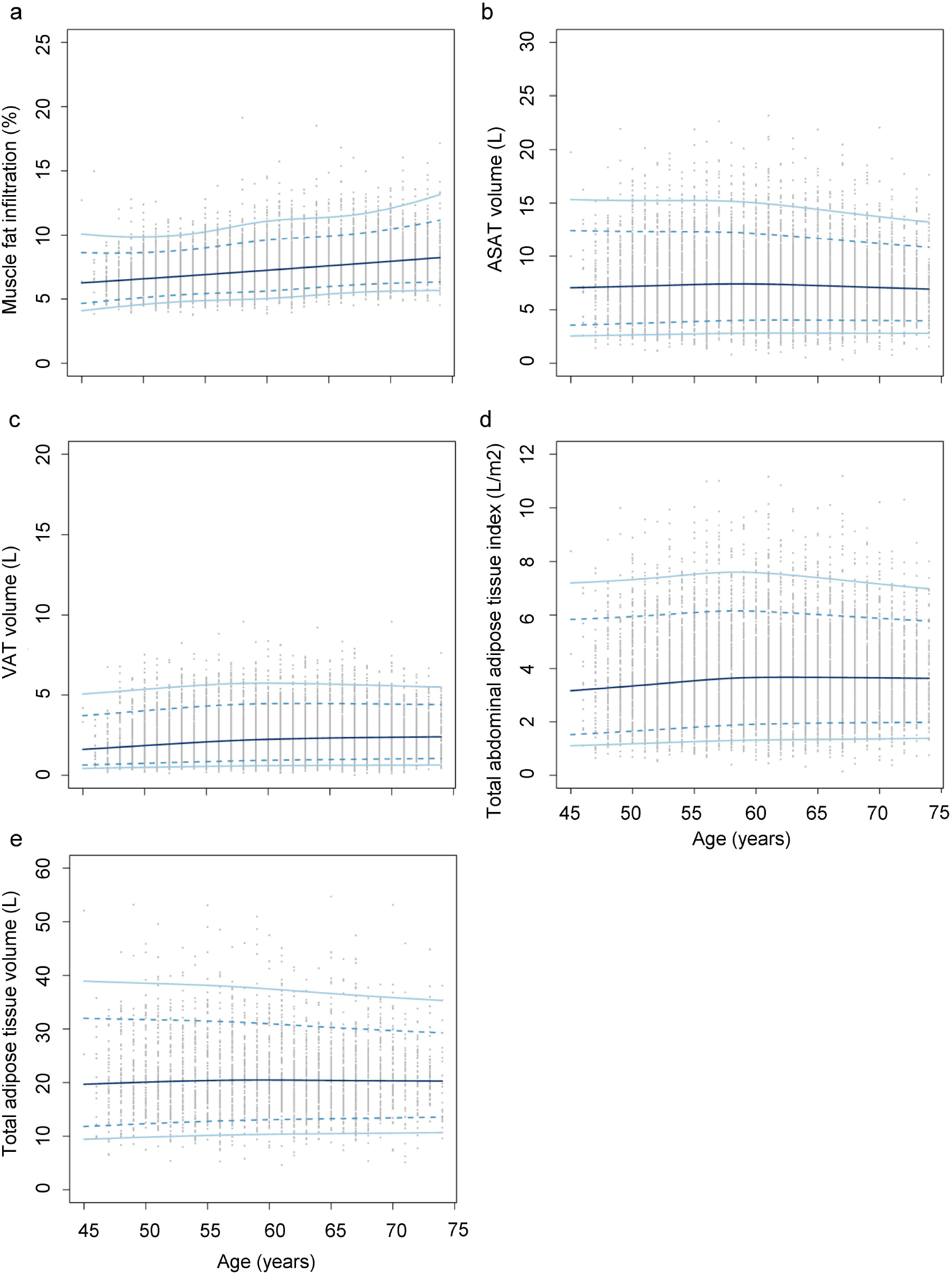
Age-specific and sex-specific percentile curves for (*a*) muscle fat infiltration, (*b*) abdominal subcutaneous adipose tissue (ASAT) volume, (*c*) visceral adipose tissue (VAT) volume, (*d*) total abdominal adipose tissue index, and (*e*) total adipose tissue volume in female.

Three levels of reference ranges for fat volume parameters are shown in *Table* 4, which were also equivalent to mean ± 1 SD, mean ± 2 SDs, and mean ± 2.5 SDs. To be noticed, lower limits of certain parameters were below zero due to modeling computation. The reference ranges for those parameters should be treated as unilateral in consideration of clinical practice. Student’s t tests between two populations showed inconsistent results among seven fat volume parameters (*Table* S1). Reference ranges for total trunk fat volume, total adipose tissue volume, total abdominal adipose tissue index in men, and ASAT volume in women may be combined between middle-aged adults and young elders. Nevertheless, based on population integrity and clinical application, we preferred separation of reference data according to the age segmentation criteria.

Reference values used in LMS method for lean volume parameters are available in Data S1.

## Discussion

Using MRI data from the UK Biobank, we have established sex-specific abdominal body composition reference ranges for middle-aged adults and young elders in a natural population cohort. To our knowledge, this is first study regarding reference development of MRI-measured body composition parameters using such a large sample size. As the golden standard of body composition assessment, the MRI reference data generated from this study can help researchers determine more accurate diagnostic criteria for overweight/obesity, sarcopenia, amyotrophy, and other metabolic disorders. The introduction of age segmentation makes the reference ranges more practical and accurate in clinical practice. In addition, the established reference ranges may also benefit renovation of classic disease risk prediction models to achieve better efficiency.

Recent studies have reported the reference ranges of body composition parameters evaluated by BIA and DXA in various countries and regions [19, 28-30]. Due to different assessment tools and measurement units, the results from this study cannot directly compared with previous findings. However, we can still roughly compare the age-related trends of similar parameters. For lean parameters, the trajectories of our results were consistent with previous studies in Caucasians [33, 36], namely lean tissue contents gradually decreased with aging. Interestingly, the fat volume parameters did not follow the trends of recent reports. Though VAT volume and muscle fat infiltration showed slight elevation with aging, other parameters kept relatively stable or presented a slight reduction in the prescribed age range (45-74 years). Accord to a previous study also using UK Biobank data [28], the fat volume index assessed by BIA presented gradual rising trend with aging. We speculated that this discrepancy may result from change of adipose density. It has been well-acknowledged that human adipose tissue composes of two main categories, namely white adipose tissue (WAT) and brown adipose tissue (BAT) [31, S7]. Based on recent researches, though the total adipose tissue volume may keep balance before extreme old age, the contents of sub-categories and fat distribution are in constant changing [32, 33]. Increasing with ages, both number and volume of WAT adipocytes increases, whereas BAT declines. In the meanwhile, ectopic fat infiltration in muscles, liver and pancreas increases. Our results have demonstrated that VAT volume and muscle fat infiltration presented gradual elevation with aging. Hence, we have reasons to speculate that, beneath the stable volume of adipose tissue, the composition of adipose tissue may have been altered, and further lead to the increasing adipose mass.

BMI and WC are two fundamental parameters for obesity diagnosis. However, based on the definition of obesity, which is characterized by an increase of body fat stores [34], simple anthropometric indices like BMI and WC apparently do not suffice. Moreover, concepts such as visceral obesity and localized adiposity gain increasing attention in recent years. Detailed assessment of adipose tissue distribution is necessary in clinical practice. DXA, a routine tool in quantifying body composition, can provide specific data on lean and fat volume of human body [35]. Whereas, as a two-dimensional scanning technique, DXA cannot distinguish visceral, deep, and superficial fat volume. MRI is a kind of three-dimensional imaging technique. The introduction of vertical axis makes it possible to obtain fat parameters of different body parts, achieving more accurate assessment. Nonetheless, medical expenditure for MRI is significantly higher than DXA. Clinicians should determine the most appropriate tool based on medical purpose and patient needs. Sarcopenia, defined as age-related loss of skeletal muscle, has driven great attention in recent decades [36, S8]. Sarcopenia is not only related to various adverse health outcomes, but dramatically increases medical expenditure [S9]. As a definite threat to public health, simple and accurate diagnostic method is particularly important for clinical practice. Currently, three parameters are evaluated for sarcopenia diagnosis, namely skeletal muscle strength, skeletal muscle mass, and physical performance [15, 37]. Among these, muscle quantity is the confirmatory index. Indeed, MRI technique serves as gold standard for non-invasive evaluation of muscle mass, while for disease diagnosis, it does not show superiority than BIA or DXA because cost-effectiveness is priorly considered and high precision is not necessary in routine practice. Therefore, though we provided reference ranges for MRI-based body composition parameters, it is still too aggressive for routine application of MRI in daily sarcopenia diagnosis.

Nevertheless, MRI technique should not be laid aside and neglected in body composition assessment. Apart from routine screening, MRI technique is more suitable for refined evaluation. Muscle quality, a relatively new concept, indicates that changes in muscle architecture and composition would impact muscle function [38], and currently can be directly assessed through highly-sensitive imaging instruments, i.e. CT and MRI [S10]. Skeletal muscle fat infiltration, also known as myosteatosis, is the main indicator for muscle quality. It has been recognized as an independent risk factor for muscle mass, strength, and mobility after adjusting other body composition parameters [39]. Moreover, the accumulation of fat in muscle would negatively impact body metabolic status, and may further link to elevated risk of various health consequences such as diabetes, fracture, late recovery and mortality [S11-12]. Due to the synergetic effect of sarcopenia and myosteatosis, it is a growing recognition that muscle mass and quality should be evaluated simultaneously in determining risk stratification in aging population. Then, as a solution, MRI technique would be an appropriate choice. We suggest that, if conditions permit, MRI examination should be arranged for individuals who are suspicious with sarcopenia and desired for detailed health instructions.

The findings in this study filled the blank of MRI-based body composition reference data. Three levels of ranges were calculated according to conventional rules. In consideration of MRI accuracy and the large sample size, we tended to recommend more rigorous reference ranges. Future researchers can consider applying these reference values as grouping criteria, then design longitudinal studies to explore the relationship between body composition and health related outcomes. Once the data accumulation is sufficient, we can amend and finalize the reference ranges of body composition parameters.

Our study had certain inherent limitations that would influence the usage of established reference ranges. All MRI-based body composition data were from the UK Biobank cohort, which only contained aged above 45 years old. The estimation of reference values for whole population cannot be implemented, and further study should be conducted to fill the blank. In addition, due to the limited sample size of ethnic minorities, the reference ranges may be only suitable for European Caucasians. Such studies should also be supplemented in other regions. Besides, our study was a cross-sectional design. No longitudinal data was involved. The age-based curves of body composition parameters were actual the single timepoint data, thus cannot be strictly defined as intra-individual trajectories. The loss of longitudinal characters may further introduce detection bias in our estimation. Currently, the UK Biobank is conducting the repeated imaging tests. When data are sufficient, the reference ranges can be adjusted to get better accuracy.

## Conclusion

In summary, we have established reference ranges for MRI-measured body composition parameters based on the UK Biobank study. The data would provide solid evidences for researchers and doctors of defining abnormal adipose and muscle conditions in middle-aged population and younger elders. Further studies are needed to supplement reference data on other age groups and ethnicities, and explore their relationship with health consequences.

## Supporting information

STROBE checklist

## Data Availability

All data produced in the present study are available upon reasonable request to the authors.

## Acknowledgements

This research was funded by the San-Ming Project of Medicine, Shenzhen (No. SZSM201812097), the Shenzhen Science and Technology Innovation Commission (No. JCYJ20200109140412476), and Peking University Shenzhen Hospital (No. LCYJ2020001).

## Ethical statement

The authors certify that they comply with the ethical guidelines for publishing in the Journal of Cachexia, Sarcopenia and Muscle: update 2021 [40].

