## Supplemental material for "Defining reference values for body composition indexes by magnetic resonance imaging in UK Biobank"

| | **Total lean tissue volume** | **Total thigh fat-free muscle volume** | **Weight-to-muscle ratio** | **Muscle fat infiltration** | **Abdominal subcutaneous adipose tissue volume** | **Visceral adipose tissue volume** | **Total trunk fat volume** | **Total adipose tissue volume** | **Total abdominal adipose tissue index** | **Abdominal fat ratio** |
|---|---|---|---|---|---|---|---|---|---|---|
| Data available (n) | 8 522 | 24 483 | 24 482 | 24 414 | 25 092 | 25 103 | 25 090 | 8 522 | 25 090 | 24 446 |
| **Exclusion based on inclusion criteria** | | | | | | | | | | |
| Total removed based on inclusion criteria (N) | 251 | 1 253 | 1 253 | 1 250 | 1 271 | 1 272 | 1 271 | 251 | 1 271 | 1 250 |
| Age criteria unmet (n) | 251 | 1 253 | 1 253 | 1 250 | 1 271 | 1 272 | 1 271 | 251 | 1 271 | 1 250 |
| Withdrawal (n) | 0 | 0 | 0 | 0 | 0 | 0 | 0 | 0 | 0 | 0 |
| **Exclusion based on exclusion criteria** | | | | | | | | | | |
| Total removed based on exclusion criteria (N) | 3 429 | 10 117 | 10 117 | 10 102 | 9 784 | 9 683 | 10 822 | 3 429 | 9 793 | 10 199 |
| Ethnic minorities (n) | 258 | 664 | 664 | 663 | 678 | 678 | 678 | 258 | 678 | 663 |
| Endocrine, nutritional or metabolic diseases (n) | 885 | 2 696 | 2 696 | 2 692 | 2 780 | 2 785 | 2 780 | 885 | 2 780 | 2 689 |
| Malignant or undetermined tumor (n) | 1 114 | 2 904 | 2 904 | 2 900 | 2 975 | 2 975 | 2 974 | 1 114 | 2 974 | 2 901 |
| HIV infection (n) | 6 | 8 | 8 | 8 | 8 | 8 | 8 | 6 | 8 | 8 |
| Medications (n) | 2 366 | 6 353 | 6 353 | 6 339 | 6 526 | 6 533 | 6 526 | 2 366 | 6 526 | 6 337 |
| Certain surgeries (n) | 35 | 94 | 94 | 93 | 94 | 94 | 94 | 35 | 94 | 94 |
| Pregnancy (n) | 0 | 0 | 0 | 0 | 0 | 0 | 0 | 0 | 0 | 0 |
| Imaging errors (n) | 3 | 1 218 | 1 218 | 1 225 | 365 | 90 | 2 071 | 3 | 383 | 1 448 |
| **Inclusion** | | | | | | | | | | |
| Analyzed (n) | 4 842 | 13 113 | 13 112 | 13 062 | 14 037 | 14 148 | 12 997 | 4 842 | 14 026 | 12 997 |

**Figure S1** Participants filtration for body composition parameters

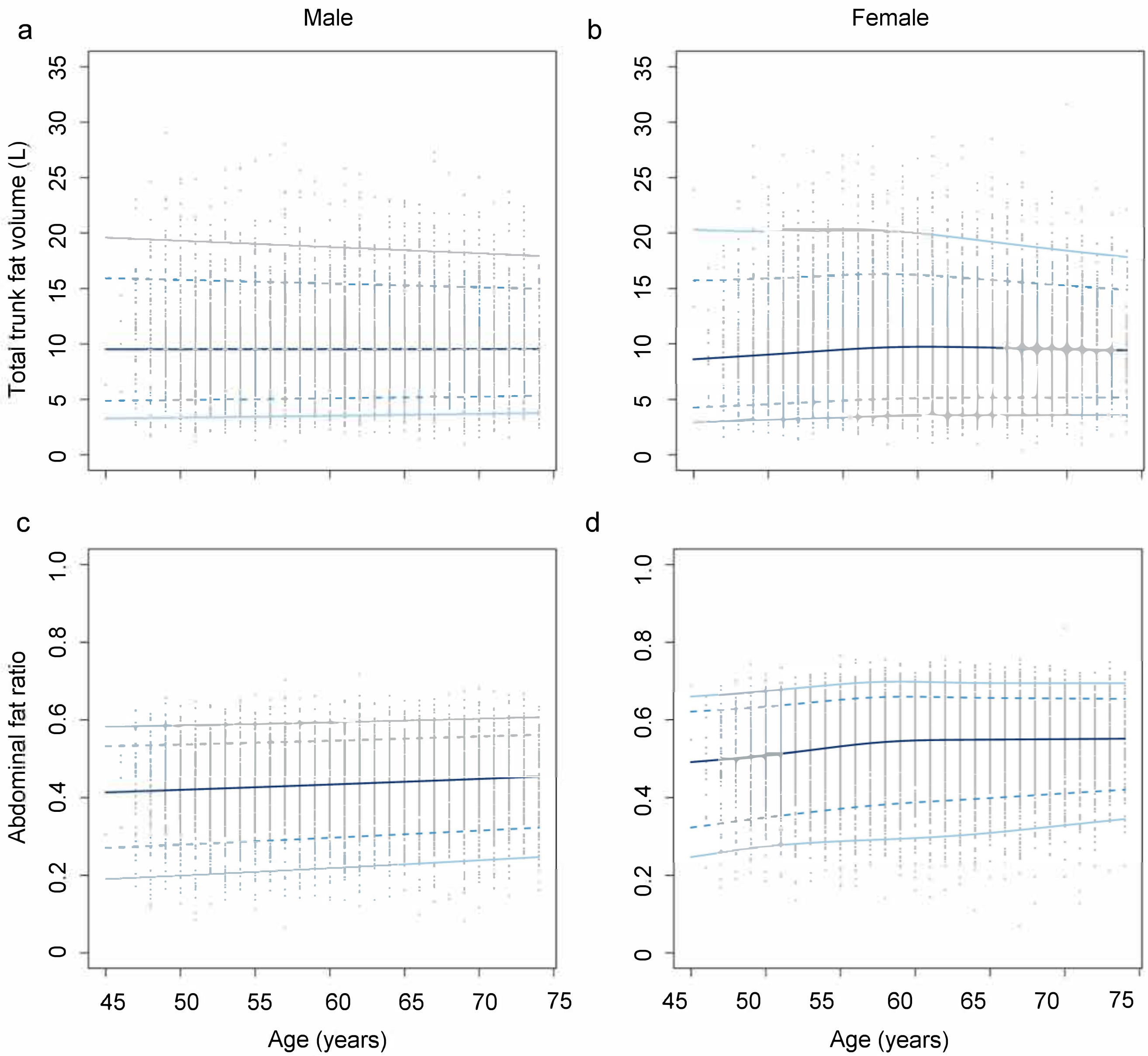


**Figure S2** Age-specific and sex-specific percentile curves for (a) total trunk fat volume in male, (b) total trunk fat volume in female, (c) abdominal fat ratio in male, and (d) abdominal fat ratio in female
